# PFAS exposure and neuroimmune and Alzheimer’s Disease–related plasma biomarkers in a rural, cognitively unimpaired population: a pilot study

**DOI:** 10.64898/2026.05.23.26353843

**Authors:** Juliana N Souza-Talarico, Hans-Joachim Lehmler, Xueshu Li, Marco Hefti, Yuxin Fun, Alaa Harb, Maria Hein, Linlin Ding, Yelena Perkhouncova

## Abstract

**INTRODUCTION:** Alzheimer’s disease (AD) is a multifactorial disorder, yet current research largely focuses on downstream biomarkers with limited attention to environmental contributors. Experimental studies suggest that per- and polyfluoroalkyl substances (PFAS) may contribute to neuroimmune and neurodegenerative pathways relevant to AD.

**OBJECTIVE:** To examine associations between PFAS exposure and neuroimmune and AD–related plasma biomarkers in cognitively unimpaired rural adults.

**METHODS:** In a cross-sectional pilot study (n=48), serum concentrations of 33 PFAS were measured, including four legacy compounds (PFOS, PFHxS, PFOA, PFNA). Plasma neuroimmune-related (ITGB2, SMOC1, TREM2, GFAP) and AD-related biomarkers (Aβ42/40, p-tau217) were detected using proteomic analysis.

**RESULTS:** PFOS showed moderate associations with ITGB2, SMOC1, and Aβ42/40 in unadjusted analyses, which attenuated after adjustment for age. PFOA and PFNA demonstrated consistent inverse associations with TREM2 before and after adjustment.

**DISCUSSION:** Findings suggest possible compound-specific PFAS associations with immune and amyloid-related biomarkers, supporting further investigation in longitudinal and mixture-based studies.

## 1. INTRODUCTION

Alzheimer’s disease (AD) is a leading cause of dementia, disability, loss of independence, and health care burden among older adults (Alzheimer’s Association, 2025). Importantly, AD develops over a prolonged preclinical phase during which biological vulnerability accumulates years to decades before the onset of clinical symptoms (Jack et al., 2013). This extended preclinical window provides a critical opportunity for prevention-oriented research to identify modifiable upstream factors that may shape cognitive vulnerability prior to clinical impairment, particularly among cognitively unimpaired middle-aged and older adults.

Despite major advances in early detection, current approaches to modeling preclinical AD risk predominantly emphasize downstream biological indicators, including genetic susceptibility (e.g., APOE ε4), neuroimaging markers, and cerebrospinal fluid or plasma biomarkers of amyloid-β (Aβ) and tau pathology (Dansson et al., 2021; Jack & Holtzman, 2013; Ossenkoppele et al., 2022). While these markers are critical for disease staging, they provide limited insight into upstream, potentially modifiable exposures and biological regulatory systems that may influence neural vulnerability across the life course, including among individuals with emerging AD-related pathology.

Persistent organic pollutants, particularly per- and polyfluoroalkyl substances (PFAS), represent a high-priority class of environmental exposures for prevention-focused brain health research (Gardener et al., 2025). PFAS are characterized by extreme environmental persistence, widespread exposure sources, and bioaccumulation in human tissues (Agency for Toxic Substances and Disease Registry [ATSDR], 2021). Data from the National Health and Nutrition Examination Survey (NHANES, 1999–2020) indicate that over 95% of the U.S. population has detectable blood levels of PFAS, including long-chain compounds such as PFOS, PFHxS, PFOA, and PFNA (Botelho et al., 2025). These long-chain (“legacy”) PFAS bind avidly to serum and tissue proteins and exhibit multi-year biological half-lives, resulting in cumulative internal exposure across adulthood and aging (ATSDR, 2021). Human exposure occurs through multiple pathways, including contaminated drinking water, food systems, consumer products, and indoor environments. Rural communities may experience heightened exposure variability due to reliance on private wells, limited regulatory monitoring infrastructure, and locally sourced food systems (EPA, 2025; Levin et al., 2024), positioning PFAS as a ubiquitous yet potentially modifiable environmental factor relevant to population-level brain health.

Accumulating experimental and human evidence supports the biological plausibility of associations between PFAS exposure and brain aging, particularly through alterations in tightly coupled neuroimmune regulatory systems implicated in neurodegenerative vulnerability (Gardener et al., 2025; Jauregui-Huerta et al., 2010). Experimental studies, largely conducted in rodents and in vitro models, demonstrate that PFAS can cross the blood–brain barrier, accumulate in brain regions central to neuroimmune and cognitive regulation—such as the hypothalamus and hippocampus—and influence glial and innate immune signaling pathways (ATSDR, 2021; Austin et al., 2003; Johansson et al., 2008; Slotkin et al., 2008; Zhang et al., 2015). For example, PFOS has been shown to accumulate in the hypothalamus at concentrations exceeding those observed in peripheral tissues (Austin et al., 2003), a region integral to immune–neuroendocrine regulation.

Across experimental models, PFAS exposure has been associated with astrocyte activation, increased expression of glial fibrillary acidic protein (GFAP), and perturbations of innate immune signaling pathways relevant to microglial function (DeWitt et al., 2012; Johansson et al., 2008; Qazi et al., 2013; Slotkin et al., 2008; Zhang et al., 2015). In parallel, human biomarker studies demonstrate that circulating GFAP increases during preclinical stages of AD, and triggering receptor expressed on myeloid cells 2 (TREM2)-related pathways play central roles in microglial responses and neurodegenerative vulnerability (Ashton et al., 2021; Benedet et al., 2021; Ulrich et al., 2017; Jin et al., 2025; Torrealba-Acosta et al., 2026).

Converging evidence further implicates PFAS exposure in processes relevant to neurodegeneration. Experimental studies report PFAS-associated alterations in hippocampal neurotransmission, catecholaminergic signaling, neuronal viability, synaptic plasticity, and learning and memory (Basaly et al., 2021; Long et al., 2013). In vitro and cerebral organoid models demonstrate PFAS-related amyloid-β (Aβ) accumulation, tau phosphorylation (p-tau), and lipid dysregulation (Lu et al., 2024; Running et al., 2024). At the population level, epidemiologic studies, including analyses of NHANES data, have reported associations between circulating PFAS concentrations and cognitive outcomes in middle-aged and older adults (Park et al., 2021; Pan et al., 2026), while emerging longitudinal studies suggest potential relationships with vascular and metabolic contributors to cognitive decline (Gardener et al., 2025). Recent detection of PFAS compounds in cerebrospinal fluid among individuals with AD biomarkers and cognitive impairment further supports central nervous system relevance in human exposure contexts (Delcourt et al., 2024).

Collectively, evidence spanning molecular, cellular, animal, and human observational domains supports the biological plausibility of associations between PFAS exposure and neuroimmune and neurodegenerative processes. However, important gaps remain in understanding whether neuroimmune alterations identified in experimental systems are observable in real-world human exposure settings, particularly during the preclinical phase of cognitive aging among cognitively unimpaired individuals. Specifically, few human studies have examined PFAS in relation to peripheral and central nervous system–related immune markers alongside AD-relevant biomarkers within the same preclinical population.

Guided by a neuroexposome framework, which emphasizes how cumulative and interacting environmental exposures across the life course become biologically embedded and shape neural vulnerability prior to overt clinical disease (Tamiz et al., 2022), this pilot study examines associations between PFAS exposure and peripheral and central nervous system–related immune markers, as well as AD–related biomarkers, in cognitively unimpaired middle-aged and older adults residing in rural communities. By focusing on early biological correlates in a real-world exposure context, this study aims to generate preliminary, hypothesis-generating evidence to inform future mechanistic and longitudinal investigations of environmental contributions to cognitive vulnerability.

## 2. METHODS

### 2.1. Study Design

This study was a cross-sectional, quantitative pilot investigation designed to examine associations between individual-level PFAS exposure and neuroimmune and AD–related plasma biomarkers in cognitively unimpaired middle-aged and older adults.

### 2.2 Study Setting and Recruitment

The study was conducted in rural communities across two counties in Eastern Iowa, Louisa County and Iowa County, located in the Midwest of the U.S. The study employed a community-embedded, decentralized research model rather than a traditional academic center–based approach, traveling to rural communities and partnering with trusted local organizations to enhance accessibility and participation. Recruitment and data collection were conducted in collaboration with community partners, including a public library, a university-affiliated primary care clinic, and a Latino community-based organization, which provided accessible venues for study activities and guidance to ensure that recruitment materials and procedures were culturally and contextually appropriate for rural populations.

Participant recruitment occurred over a 45-day period from November 1 through December 15, 2025, and reached individuals residing in seven rural communities across two counties. These communities are located near waterways with documented PFAS concentrations at or above advisory thresholds (Iowa Department of Natural Resources [IDNR]), supporting the environmental relevance of the study population and the rationale for community-based recruitment. Recruitment strategies combined community-level and individual-level outreach approaches, including in-person community tabling events, collaboration with local media outlets, flyer distribution through local businesses, and dissemination via partner organizations’ social media platforms.

### 2.3. Participants and ethical considerations

Participants were identified and enrolled using a community-engaged, non-probability sampling approach conducted on a rolling basis throughout the recruitment period. Eligible participants were community-dwelling adults aged 45–76 years, cognitively unimpaired, living independently, able to read and speak English or Spanish, and able to provide informed consent without assistance. Cognitive eligibility was confirmed using the Mini-Cog; individuals with scores ≤ 2 were excluded. Additional exclusion criteria included a self-reported diagnosis of mild cognitive impairment or dementia, major psychiatric disorders, including major depressive disorder or schizophrenia. A total of 67 individuals were recruited; 18 were excluded for not meeting eligibility criteria, and one withdrew, resulting in a final analytic sample of 48 participants. The study protocol was approved by the Institutional Review Board (#202412177). All participants provided written informed consent prior to participation.

### 2.4. Study Procedures and Data Collection

Following written informed consent, data collection was conducted using a mobile, community-based model that integrated questionnaire administration and biospecimen collection. Participants completed standardized questionnaires assessing sociodemographic characteristics, health conditions, and PFAS exposure context, followed by venous blood collection for PFAS quantification and neurobiological biomarker assessment. Blood samples were collected by licensed personnel using standardized phlebotomy procedures at community-based sites, centrifuged within 60 minutes of collection to isolate serum and plasma, aliquoted into pre-labeled polypropylene cryovials, and stored at −80 °C until analysis. All procedures were conducted by trained research personnel, including bilingual staff, who followed standardized operating manuals, with centralized coordination by the Project Coordinator (JE) and oversight by the Principal Investigator (JNST) to ensure protocol fidelity and data quality.

### 2.5. PFAS Exposure Assessment

Serum concentrations of 33 PFAS compounds were quantified using a validated liquid chromatography–tandem mass spectrometry (LC–MS/MS) method to comprehensively characterize internal PFAS exposure. Compound-specific limits of detection (LOD) ranged from 0.011 to 0.032 ng/g. Given the pilot design and modest sample size, association analyses were planned to focus on PFAS compounds demonstrating high detection frequency (≥ 90%), to support stable estimation and minimize reliance on imputation and distributional assumptions. To characterize exposure context, participants completed a PFAS exposure questionnaire adapted from ATSDR and EPA assessment tools, assessing drinking water source, receipt of contamination notices, and residential duration; these variables were summarized descriptively.

### 2.6. Outcome Neuroimmune and AD-Related Biomarkers

Plasma samples from a subset of participants were assayed using the Olink Neurodegeneration panel to quantify biomarkers reflecting neuroimmune-relevant peripheral immune activity and central nervous system–related processes (integrin beta-2 [ITGB2], SPARC-related modular calcium-binding protein 1 [SMOC1], triggering receptor expressed on myeloid cells-2 [TREM2], and glial fibrillary acidic protein [GFAP]), as well as AD disease–related biomarkers (plasma Aβ42/40 ratio and phosphorylated tau-217). These biomarkers were selected a priori based on biological relevance to neuroimmune and AD-related pathways and sufficient data completeness for exploratory analyses. Assays were conducted using Olink proximity extension assay (PEA) technology; the analyzed subset reflects the maximum sample capacity per assay batch. All samples met manufacturer-specified quality-control criteria and exhibited high detection frequency across the selected biomarkers (92.5–100%). Biomarker concentrations are reported as normalized protein expression (NPX) values, representing relative protein abundance on a log2 scale.

### 2.7. Additional variables

Additional variables, including demographic characteristics (age, sex, race, and ethnicity) and self-reported health conditions (e.g., hypertension and diabetes), were collected to characterize the study population. Age in years was selected *a priori* as the primary covariate for adjusted analyses due to its strong and well-established associations with PFAS bioaccumulation and neuroimmune and AD-related biomarker levels (Lin et al., 2025; Lu et al., 2024).

### 2.8. Statistical Analysis

All statistical analyses were conducted using SAS 9.4 (SAS Institute Inc.). Descriptive statistics were calculated to summarize participant characteristics, serum PFAS concentrations, and neuroimmune and AD-related biomarker distributions. Variable distributions were examined using graphical and analytic methods to assess normality and identify outliers. Serum PFAS concentrations were natural log-transformed for correlation and regression analyses to address right-skewed distributions, while original, untransformed PFAS concentrations were reported descriptively. In accordance with CDC/NHANES analytic guidelines, values below the limit of detection (LOD) were imputed using LOD/√2 prior to transformation and analysis (Botelho et al., 2025; Hornung et al., 1990). Using a conservative approach appropriate for a pilot study, imputation was restricted to PFAS compounds with high detection frequency (≥ 90%), while compounds with lower detection frequency were excluded from correlation and regression analyses to avoid reliance on extensive imputation and unstable estimates. Associations between log-transformed PFAS compounds with ≥ 90% detection frequency and neuroimmune and AD–related biomarker outcomes were evaluated using unadjusted Pearson correlations, age-adjusted partial correlations, and standardized linear regression coefficients adjusted for age only. The estimates are reported with corresponding 95% confidence intervals; confidence intervals for statistically significant estimates (α < 0.05) do not include zero. Pairwise correlations among PFAS compounds were computed to characterize co-exposure patterns and assess potential collinearity. Given the pilot design and modest sample size, no correction for multiple comparisons was applied, and results were interpreted as hypothesis-generating, with emphasis on effect size magnitude and direction rather than causal inference.

## 3. RESULTS

### 3.1. Participants’ characteristics, PFAS exposure, and neurobiological markers

Table 1 summarizes participant characteristics, PFAS exposures, and neurobiological marker distributions. The sample consisted primarily of older, cognitively unimpaired adults (mean age = 63.9 years, SD = 9.4 years), and was predominantly female (85.1%) and White (72.1%), with a high prevalence of cardiometabolic conditions (44.7% had hypertension and 21.7% had diabetes). On average, participants had 13.0 years of education (SD=3.7) (Table 1).

**Table 1.**
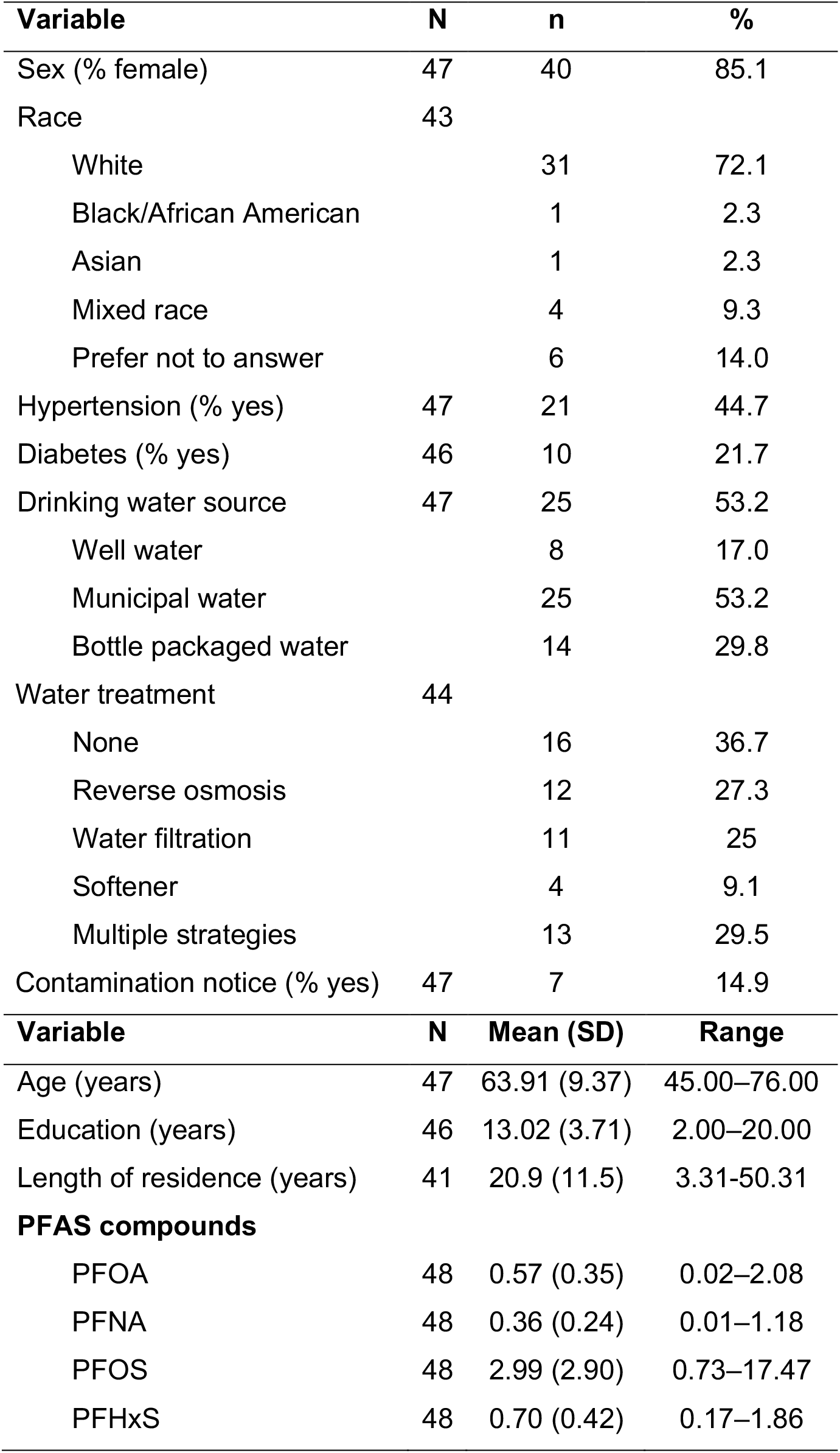
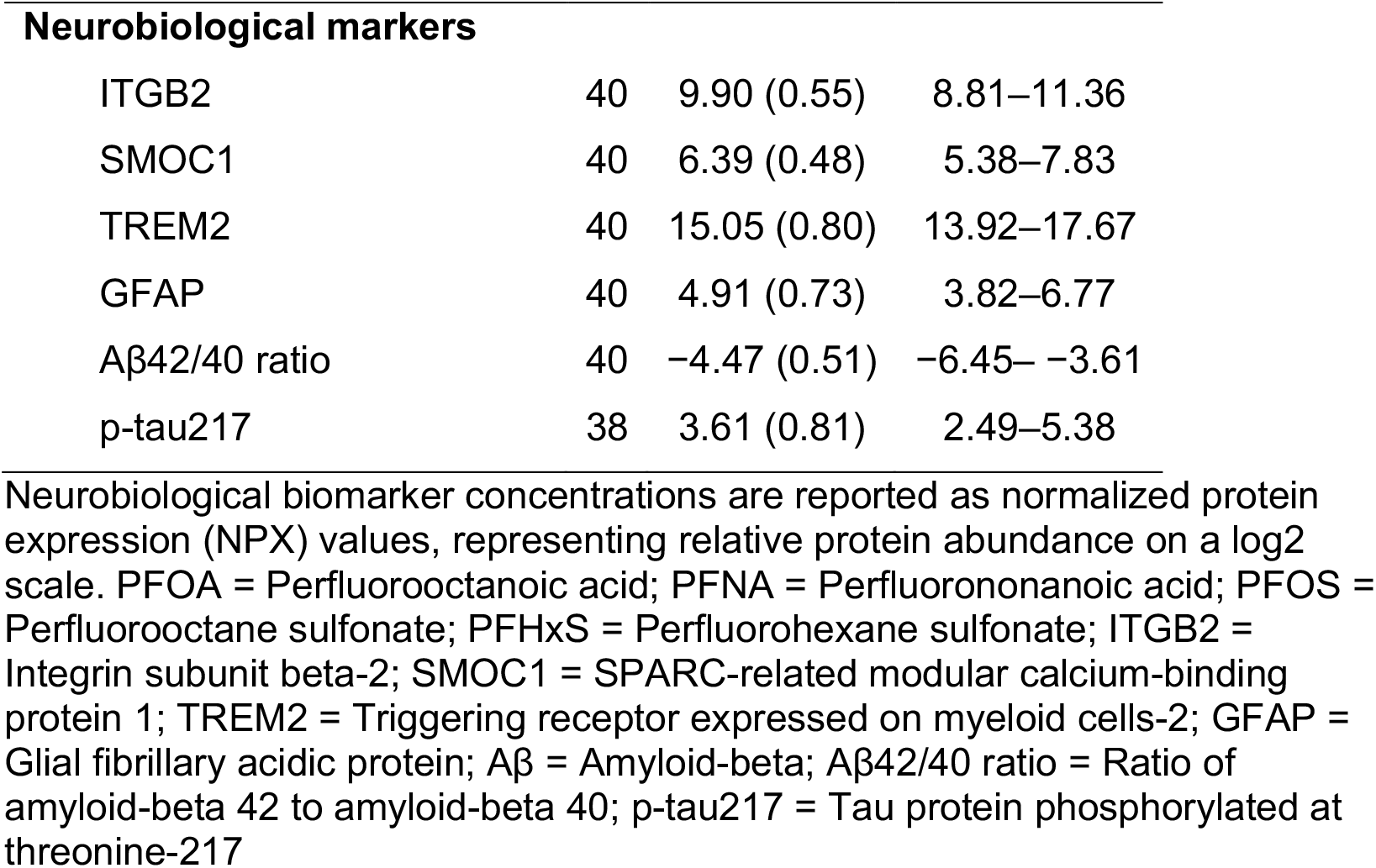
Participant Characteristics, PFAS Compounds, and Neurobiological Markers.

| **Variable** | **N** | **n** | **%** |
| --- | --- | --- | --- |
| Sex (% female) | 47 | 40 | 85.1 |
| Race | 43 |  |  |
| White |  | 31 | 72.1 |
| Black/African American |  | 1 | 2.3 |
| Asian |  | 1 | 2.3 |
| Mixed race |  | 4 | 9.3 |
| Prefer not to answer |  | 6 | 14.0 |
| Hypertension (% yes) | 47 | 21 | 44.7 |
| Diabetes (% yes) | 46 | 10 | 21.7 |
| Drinking water source | 47 | 25 | 53.2 |
| Well water |  | 8 | 17.0 |
| Municipal water |  | 25 | 53.2 |
| Bottle packaged water |  | 14 | 29.8 |
| Water treatment | 44 |  |  |
| None |  | 16 | 36.7 |
| Reverse osmosis |  | 12 | 27.3 |
| Water filtration |  | 11 | 25 |
| Softener |  | 4 | 9.1 |
| Multiple strategies |  | 13 | 29.5 |
| Contamination notice (% yes) | 47 | 7 | 14.9 |
| **Variable** | **N** | **Mean (SD)** | **Range** |
| Age (years) | 47 | 63.91 (9.37) | 45.00–76.00 |
| Education (years) | 46 | 13.02 (3.71) | 2.00–20.00 |
| Length of residence (years) | 41 | 20.9 (11.5) | 3.31–50.31 |
| **PFAS compounds** |  |  |  |
| PFOA | 48 | 0.57 (0.35) | 0.02–2.08 |
| PFNA | 48 | 0.36 (0.24) | 0.01–1.18 |
| PFOS | 48 | 2.99 (2.90) | 0.73–17.47 |
| PFHxS | 48 | 0.70 (0.42) | 0.17–1.86 |
| **Neurobiological markers** |  |  |  |
| ITGB2 | 40 | 9.90 (0.55) | 8.81–11.36 |
| SMOC1 | 40 | 6.39 (0.48) | 5.38–7.83 |
| TREM2 | 40 | 15.05 (0.80) | 13.92–17.67 |
| GFAP | 40 | 4.91 (0.73) | 3.82–6.77 |
| Aβ42/40 ratio | 40 | -4.47 (0.51) | -6.45– -3.61 |
| p-tau217 | 38 | 3.61 (0.81) | 2.49–5.38 |

Neurobiological biomarker concentrations are reported as normalized protein expression (NPX) values, representing relative protein abundance on a log2 scale. PFOA = Perfluorooctanoic acid; PFNA = Perfluorononanoic acid; PFOS = Perfluorooctane sulfonate; PFHxS = Perfluorohexane sulfonate; ITGB2 = Integrin subunit beta-2; SMOC1 = SPARC-related modular calcium-binding protein 1; TREM2 = Triggering receptor expressed on myeloid cells-2; GFAP = Glial fibrillary acidic protein; Aβ = Amyloid-beta; Aβ42/40 ratio = Ratio of amyloid-beta 42 to amyloid-beta 40; p-tau217 = Tau protein phosphorylated at threonine-217

Detection frequency varied substantially across PFAS compounds. Among the 33 PFAS included in the analytical panel, the exposure profile presented in the main results was dominated by four legacy PFAS (PFOS, PFHxS, PFNA, and PFOA) with high detection frequency. PFHxS and PFOS were detected in all samples, followed by PFNA (93.8%) and PFOA (91.7%), which were detected in most samples. Among these compounds, PFOS demonstrated the greatest variability in measured concentrations (SD=2.90), consistent with heterogeneous exposure levels within the study population (Table 1). In contrast, several perfluorocarboxylic acids, sulfonates, and fluorotelomer compounds were detected in fewer than half of samples, and multiple compounds were not detected in any sample, including PFBA, PFHxA, PFBS, PFNS, PFDoS, 4:2 FTS, HFPO-DA, 11Cl-PF3OUdS, FBSA, FHxSA, FOSA, and NaDONA. Descriptive statistics for all PFAS measured, including compounds with lower detection frequency, are provided in the Supplementary Materials (Table S1) to ensure transparent exposure characterization.

Regarding exposure context, participants reported using a mix of drinking water sources, including municipal (53.2%), well (17.0%), and bottled water (29.8%), and some (14.9%) reported receiving a contamination notice for their household water supply (Table 1). Among participants using well water, the average duration of well use was approximately 5.1 years (range: 4–6 years). Participants also demonstrated substantial residential stability, with long durations (mean 20.9 years, SD=11.5) at their current residence (Table 1), indicating potential for cumulative environmental exposure. With respect to water treatment, 36.7% of participants reported no treatment; while among those who treated their water, reverse osmosis was the most reported method (27.3%) (Table 1). Moderate pairwise intercorrelations were observed among the four legacy PFAS compounds (PFOS, PFHxS, PFNA, and PFOA), indicating non-independent exposure distributions (Supplementary Table S2).

Neurobiological marker distributions are summarized in Table 1. All selected biomarkers demonstrated high detection frequency and substantial interindividual variability, supporting their inclusion in exploratory association analyses (Table 1). Additional proteins included in the Olink Neurodegeneration panel were measured but are not presented in the main table. Detection frequency and descriptive statistics for the full panel are provided in the Supplementary Materials (Table S3) to ensure transparency and comprehensive reporting.

### 3.2. Association between PFAS, Neuroimmune, and AD-related biomarkers

Based on detection frequency criteria, the primary exposures of interest for the association analyses were four legacy PFAS compounds (PFOS, PFHxS, PFOA, and PFNA), with serum concentrations log-transformed for analysis. Patterns of associations differed between unadjusted and age-adjusted analyses, with several associations attenuating after adjustment for age (Table 2).

**Table 2.** Associations Between Log-Transformed PFAS Compounds with ≥ 90% Detection frequency and Neuroimmune and AD-related Biomarkers.

| Biomarkers | Statistic | PFOA | PFNA | PFOS | PFHxS |
| --- | --- | --- | --- | --- | --- |
| ITGB2 | *r* | 0.08 (-0.23–0.38) | 0.01 (-0.30–0.32) | **0.32 (0.01–0.58)** | 0.14 (-0.18–0.43) |
|  | *r<sub>p</sub>* | 0.06 (-0.27–0.37) | -0.00 (-0.32–0.32) | 0.28 (-0.04–0.55) | 0.08 (-0.25–0.39) |
| | $\beta$ | 0.06 (-0.28–0.40) | -0.00 (-0.34–0.33) | 0.33 (-0.04–0.69) | 0.09 (-0.28–0.46) |
| SMOC1 | *r* | 0.14 (-0.18–0.43) | -0.14 (-0.43–0.18) | **0.36 (0.05–0.60)** | 0.10 (-0.22–0.40) |
|  | *r<sub>p</sub>* | 0.09 (-0.24–0.40) | -0.19 (-0.48–0.14) | 0.22 (-0.11–0.50) | -0.08 (-0.39–0.24) |
| | $\beta$ | 0.09 (-0.23–0.41) | -0.18 (-0.49–0.14) | 0.23 (-0.12–0.59) | -0.09 (-0.44–0.26) |
| TREM2 | *r* | **-0.35 (-0.59– -0.04)** | **-0.53 (-0.73– -0.27)** | -0.11 (-0.41–0.21) | -0.06 (-0.37–0.25) |
|  | *r<sub>p</sub>* | **-0.40 (-0.64– -0.09)** | **-0.58 (-0.76– -0.32)** | -0.28 (-0.55–0.05) | -0.20 (-0.49–0.13) |
| | $\beta$ | **-0.40 (-0.71– -0.10)** | **-0.57 (-0.84– -0.30)** | -0.31 (-0.67–0.05) | -0.22 (-0.58–0.14) |
| GFAP | *r* | 0.04 (-0.27–0.35) | 0.01 (-0.30–0.32) | 0.23 (-0.09–0.50) | 0.22 (-0.10–0.50) |
|  | *r<sub>p</sub>* | -0.05 (-0.36–0.27) | -0.04 (-0.36–0.28) | -0.02 (-0.33–0.30) | 0.00 (-0.32–0.32) |
| | $\beta$ | -0.05 (-0.34–0.25) | -0.04 (-0.33–0.26) | -0.02 (-0.35–0.32) | 0.00 (-0.32–0.33) |
| Aβ 42/40 ratio | *r* | -0.08 (-0.38–0.24) | -0.14 (-0.43–0.18) | **-0.37 (-0.61– -0.07)** | -0.28 (-0.54–0.04) |
|  | *r<sub>p</sub>* | -0.03 (-0.35–0.29) | -0.12 (-0.42–0.21) | -0.27 (-0.55–0.05) | -0.17 (-0.46–0.16) |
| | $\beta$ | -0.03 (-0.36–0.30) | -0.12 (-0.44–0.21) | -0.31 (-0.66–0.05) | -0.18 (-0.54–0.17) |
| p-tau217<sup>a</sup> | *r* | 0.06 (-0.26–0.38) | -0.10 (-0.40–0.23) | 0.13 (-0.20–0.43) | 0.19 (-0.14–0.48) |
|  | *r<sub>p</sub>* | 0.02 (-0.32–0.34) | -0.14 (-0.45–0.20) | -0.05 (-0.37–0.28) | 0.04 (-0.29–0.37) |
| | $\beta$ | 0.01 (-0.32–0.35) | -0.13 (-0.46–0.19) | -0.06 (-0.44–0.32) | 0.05 (-0.32–0.41) |

Note. N=40; <sup>a</sup> n=38. 95% confidence intervals for coefficient estimates are shown in parentheses; bolded values indicate that the intervals do not include zero. *r* = unadjusted Pearson correlation estimate; *r<sub>p</sub>* = age-adjusted partial correlation estimate; $\beta$ = standardized regression coefficient adjusted for age. Estimates were calculated using pair-specific complete cases; one participant with missing age was not included in age-adjusted analyses. Neurobiological biomarker concentrations are reported as normalized protein expression (NPX) values, representing relative protein abundance on a log2 scale. PFOA = Perfluorooctanoic acid; PFNA = Perfluorononanoic acid; PFOS = Perfluorooctane sulfonate; PFHxS = Perfluorohexane sulfonate; ITGB2 = Integrin subunit beta-2; SMOC1 = SPARC-related modular calcium-binding protein 1; TREM2 = Triggering receptor expressed on myeloid cells-2; GFAP = Glial fibrillary acidic protein; Aβ = Amyloid-beta; Aβ42/40 ratio = Ratio of amyloid-beta 42 to amyloid-beta 40; p-tau217 = Tau protein phosphorylated at threonine-217

In unadjusted analyses, PFOS showed moderate positive correlations with immune signaling-related biomarkers, including ITGB2 (r = 0.32) and SMOC1 (r = 0.36), and a moderate inverse correlation with the Aβ42/40 ratio (r = −0.37). After adjustment for age, associations between PFOS and ITGB2, SMOC1, and Aβ42/40 were attenuated (ITGB2: β = 0.33; SMOC1: β = 0.23; Aβ42/40: β = −0.31), suggesting confounding by age.

PFOA and PFNA exhibited modest inverse associations with TREM2 in both unadjusted and adjusted analyses (PFOA: r = −0.35 and β = −0.40; PFNA: r = −0.53 and β = -0.57), indicating that these relationships were not explained by age. A modest inverse association between PFOS and TREM2 emerged after adjustment for age (β = −0.31), although the confidence interval included zero, suggesting potential negative confounding by age.

Across all four PFAS compounds, associations with astrocytic (GFAP) and tau-related (p-tau217) biomarkers were small and near null after age adjustment (|β| < 0.15). Similarly, PFHxS showed consistently small associations with neuroimmune and AD–related biomarkers in both unadjusted and adjusted analyses.

## 4. DISCUSSION

Guided by a neural exposome framework, this pilot study examined associations between PFAS exposure and peripheral neuroimmune and AD–related plasma biomarkers in cognitively unimpaired middle-aged and older adults residing in rural communities. PFAS exposure in this cohort was dominated by widely detected legacy compounds (PFOS, PFHxS, PFOA, and PFNA), while many other PFAS were present at low or undetectable levels, reflecting heterogeneous, non-uniform exposure patterns common in rural settings. Comparison of unadjusted and age-adjusted associations indicated that several PFAS–biomarker associations, particularly those involving PFOS with ITGB2, SMOC1, and the Aβ42/40 ratio, were partially attenuated after accounting for age, whereas modest inverse associations between PFOA, PFNA, and TREM2 were preserved following age adjustment. In addition, observed intercorrelations among PFAS compounds underscore the challenge of isolating individual compound effects in real-world exposure contexts and highlight the need for larger studies capable of multi-pollutant modeling. Overall, these findings suggest that PFAS compound-specific associations observed in this study are biologically plausible and consistent with emerging experimental and epidemiologic evidence, pointing to potential links between select PFAS compounds and microglial-related immune signaling that warrant further investigation in larger, longitudinal studies.

Across analytic models, PFOS showed the most consistent associations with select neuroimmune and amyloid-related plasma biomarkers, although several associations were attenuated after adjustment for age. In unadjusted analyses, higher PFOS concentrations were moderately associated with increased levels of ITGB2 and SMOC1, biomarkers involved in immune signaling and extracellular matrix dynamics. ITGB2 encodes the β2 integrin subunit (CD18), which is expressed on leukocytes and plays a central role in immune cell adhesion, trafficking, and activation during inflammatory responses (Bednarczyk et al., 2020). Experimental studies indicate that PFOS can activate innate immune signaling pathways and promote pro-inflammatory responses, supporting the biological plausibility of PFOS-related perturbations in β2 integrin–mediated immune processes (Bednarczyk et al., 2020; Bharal et al., 2024). SMOC1 is a matricellular protein involved in extracellular matrix organization, immune-related tissue remodeling, and neuronal–glial interactions and has recently emerged as a biomarker associated with early AD pathology (Balcomb et al., 2024; Afroz et al., 2024). Proteomic and neuropathological studies demonstrate that SMOC1 is among the earliest changing proteins in AD and shows increased levels in cerebrospinal fluid during preclinical disease stages (Balcomb et al., 2024). SMOC1 has been shown to be enriched in brain regions containing amyloid plaques and to directly interact with amyloid-β in human brain tissue, influencing aggregation kinetics and reflecting early neuropathological changes (Balcomb et al., 2024). The positive association between PFOS and SMOC1 observed in this study may therefore reflect PFOS-related disruptions in immune–extracellular matrix interactions that intersect with early neurodegenerative processes. PFOS also showed a moderate inverse association with the plasma Aβ42/40 ratio in unadjusted models, a direction consistent with a peripheral biomarker pattern that has been associated with altered amyloid processing and shown to closely reflect cerebral amyloid burden in preclinical and cognitively unimpaired populations (Schindler et al., 2019; Janelidze et al., 2020; Palmqvist et al., 2020). Although this association was attenuated after age adjustment, the observed pattern aligns with experimental evidence demonstrating that PFOS exposure can influence amyloid precursor protein processing, increase amyloidogenic signaling, and alter amyloid-β metabolism in animal and cellular models (Basaly et al., 2021; Wilkins & Swerdlow, 2016).

PFOA and PFNA showed the strongest associations with TREM2, with moderate inverse relationships that were preserved after age adjustment. TREM2 is primarily expressed by microglia, but soluble TREM2 can also be measured in peripheral circulation, where it reflects immune-related signaling linked to myeloid cell activation and inflammatory regulation rather than direct measures of central nervous system pathology (Suárez-Calvet et al., 2016; Heslegrave et al., 2016). Experimental studies demonstrate that PFAS exposure, including PFOA and PFNA, can disrupt immune signaling pathways, promote oxidative stress, and alter inflammatory responses in macrophage and microglial analogs, providing biological plausibility for PFAS-related modulation of TREM2-associated immune processes (Basaly et al., 2021; Bharal et al., 2024). In addition, PFAS have been shown to affect lipid metabolism and membrane signaling domains, which are integral to TREM2 function and downstream immune regulation (Ullrich et al., 2017). Importantly, the observed inverse associations in this study were detected in plasma and in a cognitively unimpaired population, suggesting that these relationships may reflect early or systemic immune alterations rather than direct microglial dysfunction within the brain. Within the context of a pilot study and environmental (non-occupational) exposure levels, these findings suggest that PFOA and PFNA may be linked to subtle immune signaling changes relevant to microglial pathways, while underscoring the need for cautious interpretation and validation in larger studies incorporating longitudinal designs and central biomarkers.

By contrast, associations between PFAS and tau-related pathology (p-tau217) were small and near null after age adjustment across all compounds. This suggests that, in this cognitively unimpaired sample, PFAS exposure may be more closely linked to immune and amyloid-related processes than to tau phosphorylation, which is generally considered a later-stage marker of AD neurodegeneration (Palmqvist et al., 2024; Therriault et al., 2024). Similarly, PFHxS demonstrated uniformly small and inconsistent associations across neuroimmune and AD-related biomarkers, which may reflect differences in toxicokinetic, bioaccumulation, or central nervous system penetration compared with PFOS and PFOA (Cao & Ng, 2021; Song et al., 2026), or limited statistical power in this pilot study.

Importantly, the present findings extend the evidence from experimental studies by suggesting that such biologically relevant associations may also be detectable in humans experiencing typical environmental exposure, rather than only under conditions of high-dose or occupational exposure. Within the limitations of a small pilot sample, these results support the plausibility that background, chronic PFOS exposure may contribute to subtle neuroimmune and amyloid-related alterations, underscoring the need for larger, longitudinal studies to clarify dose–response relationships and temporal dynamics.

Within a neural exposome framework, these findings underscore the importance of compound-specific effects and early biological perturbations that may precede overt cognitive decline (Ta. The cohort consisted of cognitively unimpaired adults with long residential stability and varied drinking water sources, conditions under which chronic, low-level exposure may accumulate over time. Rural populations relying on private wells or small municipal systems may experience heightened or undercharacterized PFAS exposure, particularly where routine monitoring is limited (Wood et al., 2025; U.S. Geological Survey, 2023). Integrating environmental exposure assessment with sensitive plasma biomarkers may therefore be especially valuable for detecting early biological vulnerability in such populations.

The observed associations should also be considered in the context of participants’ exposure environments. In this cohort, the use of mixed drinking water sources, including municipal and well water, alongside variable adoption of household treatment systems, suggests heterogeneous but potentially sustained exposure pathways. When combined with prolonged residential stability, these factors may contribute to cumulative PFAS exposure over time. The moderate intercorrelations observed among PFAS compounds further support the presence of shared exposure pathways, consistent with drinking water–related co-exposure. Together, these findings highlight the importance of integrating real-world exposure context into the interpretation of PFAS–biomarker associations, particularly in rural settings where exposure sources may be diffuse, variable, and not routinely characterized.

Several limitations should be noted. The cross-sectional design limits causal inference, and the modest sample size constrains statistical power and precision. The study sample was predominantly female, which may limit generalizability given documented sex-dependent differences in PFAS pharmacokinetics and health associations (Jain & Ducatman, 2022; Johnson et al., 2025). Residual confounding remains possible, in part due to the limited set of covariates included in the analyses, the modest sample size, and potential influences on PFAS concentrations and neurobiological marker levels, which could affect the magnitude or direction of the observed associations. In addition, the modest sample size limited the ability to evaluate PFAS co-occurrence and mixture effects using multi-pollutant analytical approaches (e.g., the quantile g-computation approach), which require larger samples to support stable estimation in the presence of correlated exposures (Keil et al., 2020).

Despite these limitations, the study has several notable strengths. The community-embedded, decentralized research model facilitated recruitment of a geographically and socioeconomically diverse rural population that is often underrepresented in biomarker research. The use of serum PFAS concentrations provided an objective measure of internal exposure across an extensive panel of compounds, complemented by contextual environmental exposure information. In addition, the application of proteomic technology enabled sensitive quantification of neuroimmune and AD–related biomarkers. Finally, the focus on cognitively unimpaired individuals enhances the relevance of these findings for understanding early biological changes relevant to AD and prevention-oriented studies.

## 5. CONCLUSIONS AND FUTURE DIRECTIONS

In summary, this pilot study provides preliminary evidence of compound-specific associations between PFAS exposure and neuroimmune and AD–related plasma biomarkers in cognitively unimpaired adults. Patterns of effect sizes suggest that PFOS is most consistently associated with markers of immune signaling and amyloid-related plasma biomarkers, whereas PFOA and PFNA show more consistent inverse associations with TREM2, highlighting potential links to myeloid-related immune pathways. These findings should be interpreted as hypothesis-generating and suggest that distinct PFAS compounds may differentially influence neuroimmune and amyloid-related processes at levels consistent with typical environmental exposure. The moderate intercorrelations observed among PFAS further underscore the importance of considering co-occurring exposures, although the current sample size limited the ability to evaluate PFAS mixtures using multi-pollutant modeling approaches. Future research should prioritize larger, longitudinal studies that integrate mixture-based exposure frameworks with sensitive biomarkers across the neuroimmune and neurodegenerative continuum. Such approaches will be critical to clarify dose–response relationships, temporal dynamics, and the cumulative impact of environmental exposures on early neural vulnerability. Together, these findings highlight the value of a neural exposome perspective for understanding how co-occurring environmental contaminants may contribute to early biological changes preceding clinical cognitive impairment.

## Data Availability

The de-identified dataset supporting the findings of this study will be made available upon reasonable request to the corresponding author, in accordance with institutional policies, IRB approval and applicable data management and sharing requirements

## ACKNOWLEDGMENTS

The authors gratefully acknowledge the community organizations that partnered in this research and provided trusted, accessible spaces for recruitment and data collection, including local public libraries, a university-affiliated primary care clinic, and a Latino community-based organization. We also extend our sincere appreciation to the community members who participated in the study and generously contributed their time and trust.

## CONSENT STATEMENT

Participants provided written informed consent prior to enrollment in the study

