## Supplemental Materials for "PFAS exposure and neuroimmune and Alzheimer’s Disease–related plasma biomarkers in a rural, cognitively unimpaired population: a pilot study"

### **Table S1.** PFAS Serum Concentrations and Detection frequency in the Study Sample (N = 48)

| **PFAS ng/g** | **N detected** | **% detection frequency** | **Mean** | **SD** | **Minimum** | **Maximum** |
| --- | --- | --- | --- | --- | --- | --- |
| PFOS | 48 | 100.0 | 2.99 | 2.90 | 0.73 | 17.47 |
| PFHxS | 48 | 100.0 | 0.70 | 0.42 | 0.17 | 1.86 |
| PFNA | 45 | 93.8 | 0.39 | 0.23 | 0.12 | 1.18 |
| PFOA | 44 | 91.7 | 0.62 | 0.33 | 0.00 | 2.00 |
| PFHpS | 41 | 85.4 | 0.10 | 0.06 | 0.04 | 0.32 |
| NMeFOSAA | 32 | 66.7 | 0.12 | 0.15 | 0.04 | 0.82 |
| PFHxDA | 29 | 60.4 | 0.39 | 0.14 | 0.20 | 0.76 |
| PFTeDA | 26 | 54.2 | 0.84 | 0.39 | 0.37 | 1.79 |
| PFUnA | 25 | 52.1 | 0.10 | 0.06 | 0.05 | 0.31 |
| PFTrDA | 22 | 45.8 | 0.28 | 0.32 | 0.10 | 1.27 |
| PFDA | 16 | 33.3 | 0.26 | 0.17 | 0.16 | 0.70 |
| PFHpA | 7 | 14.6 | 2.74 | 0.64 | 2.15 | 4.04 |
| 6:2 FTS | 6 | 12.5 | 4.07 | 0.54 | 3.02 | 4.41 |
| 8:2 FTS | 5 | 10.4 | 0.06 | 0.03 | 0.04 | 0.11 |
| PFPeS | 4 | 8.3 | 0.03 | 0.01 | 0.02 | 0.04 |
| PFODA | 3 | 6.3 | 0.29 | 0.08 | 0.20 | 0.35 |
| PFPeA | 2 | 4.2 | 0.63 | 0.71 | 0.12 | 1.13 |
| PFDoA | 1 | 2.1 | 0.25 | — | 0.25 | 0.25 |
| PFDS | 1 | 2.1 | 0.14 | — | 0.14 | 0.14 |
| NEtFOSAA | 1 | 2.1 | 0.10 | — | 0.10 | 0.10 |
| 9Cl‑PF3ONS | 1 | 2.1 | 0.04 | — | 0.04 | 0.04 |
| PFBA | 0 | 0.0 | — | — | — | — |
| PFHxA | 0 | 0.0 | — | — | — | — |
| PFBS |  |  |  |  |  |  |
| PFNS | 0 | 0.0 | — | — | — | — |
| PFDoS |  |  |  |  |  |  |
| 4:2FTS | 0 | 0.0 | — | — | — | — |
| HFPO-DA |  |  |  |  |  |  |
| 11Cl-PF3OUds | 0 | 0.0 | — | — | — | — |
| FBSA |  |  |  |  |  |  |
| FHxSA | 0 | 0.0 | — | — | — | — |
| FOSA |  |  |  |  |  |  |
| NaDONA | 0 | 0.0 | — | — | — | — |

Percent detection frequency calculated as the number of samples with detected concentrations divided by the total number of samples analyzed (N = 48). Summary statistics are presented for detected values only. SD not calculated where N ≤ 1.

**Table S2. Pearson Correlation Estimates Among Log-Transformed PFAS Compounds with ≥ 90% Detection frequency**

| **Variable** | **ln (PFOA)** | **ln (PFNA)** | **ln (PFOS)** | **ln (PFHxS)** |
| --- | --- | --- | --- | --- |
| ln (PFOA) | - |  |  |  |
| ln (PFNA) | 0.59 (0.37–0.75) | - |  |  |
| ln (PFOS) | 0.52 (0.28–0.70) | 0.55 (0.31–0.72) | - |  |
| ln (PFHxS) | 0.62 (0.40–0.77) | 0.44 (0.18–0.64) | 0.61 (0.39–0.76) | - |

Note. N=48; 95% confidence intervals for correlation estimates are shown in parentheses.

### **Table S3**. Detection frequency and Descriptive Statistics for Neurodegeneration-related Biomarkers

| **Biomarker (NPX)** | **N detected** | **% detection frequency** | **Mean** | **SD** | **Min** | **Max** |
| --- | --- | --- | --- | --- | --- | --- |
| AGER | 40 | 100.0 | 13.28 | 0.52 | 12.19 | 14.27 |
| Aβ40 | 40 | 100.0 | 10.28 | 0.52 | 9.41 | 11.87 |
| Aβ42 | 40 | 100.0 | 5.81 | 0.66 | 4.35 | 7.83 |
| BACE1 | 40 | 100.0 | 6.79 | 0.28 | 6.16 | 7.61 |
| BMP7 | 40 | 100.0 | 6.67 | 0.28 | 5.79 | 7.35 |
| CLSTN3 | 40 | 100.0 | 4.39 | 0.41 | 3.62 | 5.35 |
| DDAH1 | 40 | 100.0 | 7.22 | 0.65 | 5.94 | 9.51 |
| DDC | 40 | 100.0 | 11.27 | 0.70 | 10.16 | 13.49 |
| EIF2AK2 | 40 | 100.0 | 12.20 | 0.77 | 9.99 | 13.81 |
| ENO2 | 40 | 100.0 | 7.80 | 0.50 | 6.68 | 8.71 |
| FOXO3 | 40 | 100.0 | 5.15 | 0.55 | 3.65 | 6.13 |
| GDNF | 32 | 80.0 | 2.61 | 0.35 | 2.18 | 3.46 |
| GFAP | 40 | 100.0 | 4.91 | 0.73 | 3.82 | 6.77 |
| GLRX | 40 | 100.0 | 8.47 | 0.49 | 7.14 | 9.52 |
| HLA‑DRA | 35 | 87.5 | 1.63 | 0.35 | 1.06 | 2.85 |
| ITGAM | 40 | 100.0 | 9.13 | 0.28 | 8.31 | 9.78 |
| ITGB2 | 40 | 100.0 | 9.90 | 0.55 | 8.81 | 11.36 |
| KLK8 | 40 | 100.0 | 9.15 | 0.56 | 8.38 | 10.93 |
| MMP10 | 40 | 100.0 | 12.00 | 0.65 | 10.05 | 13.34 |
| MOG | 40 | 100.0 | 9.15 | 0.49 | 8.24 | 10.41 |
| NEFL | 40 | 100.0 | 4.49 | 0.84 | 3.32 | 6.99 |
| NPTX1 | 40 | 100.0 | 8.37 | 0.48 | 6.96 | 9.15 |
| NPTX2 | 40 | 100.0 | 9.00 | 0.46 | 7.96 | 9.88 |
| NPTXR | 40 | 100.0 | 11.39 | 0.39 | 10.42 | 12.13 |
| OMG | 40 | 100.0 | 6.46 | 0.69 | 5.09 | 7.82 |
| RTN4R | 40 | 100.0 | 4.65 | 0.49 | 3.26 | 5.58 |
| SCG2 | 40 | 100.0 | 7.20 | 0.42 | 6.45 | 8.52 |
| SDC4 | 40 | 100.0 | 8.19 | 0.47 | 6.72 | 8.91 |
| SMOC1 | 40 | 100.0 | 6.39 | 0.48 | 5.38 | 7.83 |
| STX1B | 40 | 100.0 | 4.29 | 0.43 | 3.56 | 5.36 |
| SYT1 | 40 | 100.0 | 3.73 | 0.53 | 2.78 | 4.87 |
| TP53 | 40 | 100.0 | 5.66 | 0.66 | 4.46 | 6.82 |
| TREM1 | 40 | 100.0 | 14.10 | 0.43 | 13.36 | 15.71 |
| TREM2 | 40 | 100.0 | 15.05 | 0.80 | 13.92 | 17.67 |
| VSNL1 | 40 | 100.0 | 3.36 | 0.38 | 2.28 | 4.29 |
| WWOX | 40 | 100.0 | 3.43 | 0.90 | 1.43 | 5.21 |
| p‑tau217 | 38 | 95.0 | 3.61 | 0.81 | 2.49 | 5.38 |

Detection frequency calculated as the number of samples with valid NPX values divided by the total number of samples analyzed (N = 48). NPX values represent normalized protein expression on a log2 scale.
